# Lack of consideration of sex and gender in clinical trials for COVID-19

**DOI:** 10.1101/2020.09.13.20193680

**Authors:** Emer Brady, Mathias Wullum Nielsen, Jens Peter Andersen, Sabine Oertelt-Prigione

## Abstract

Sex and gender differences impact the incidence of SARS-CoV-2 infection and COVID-19 mortality. Furthermore, sex differences characterize the frequency and severity of pharmacological side effects. A large number of clinical trials are ongoing to develop new therapeutic approaches and vaccines for COVID-19. We investigated the inclusion of sex and/or gender in currently registered studies on http://ClinicalTrials.gov. Only 416 (16.7%) of the 2,484 registered SARS-CoV-2/COVID-19 trials mention sex/gender as recruitment criterion and only 103 (4.1%) allude to sex/gender in the description of the analysis phase. None of the 11 clinical trials published in scientific journals on June 2020 reported sex-disaggregated results. Hence, lack of consideration upon registration does not seem to be corrected during trial execution and reporting. Given the biological relevance and the potential risks of unwanted side effects, we urge researchers to focus on sex-disaggregated analyses already at the planning stage of COVID-19 trials.

## Introduction

Testing protocols, diagnostic algorithms and hospitalization criteria for SARS-CoV-2 infections and COVID-19 may vary between countries, due to differences in resources, national guidelines and phase of the pandemic. Nevertheless, available data point towards an increased risk of mortality for male patients with COVID-19 worldwide compared to female patients^1^. This could be related to intrinsic sex differences in the immune reaction^2^ or specific characteristics of the SARS-CoV-2 infectious process. The virus connects to the ACE2 receptor, which is encoded on the X chromosome and co-engages a serine protease – TMPRSS2 – that appears to be hormone-sensitive^3^. A recent report also highlights the role of the innate immune response in the fight against the virus; specifically of TLR-7^4^, which is also encoded on the X chromosome. TLR-7 has been previously described as a relevant modulator of sex-specific differences in anti-viral immunity^5^. The investigation of sex differences could provide essential insights into COVID-19 pathophysiology and possibly aid the identification of effective interventions. In addition to the study of sex differences, the analysis of the impact of gender is warranted^6^. Gender, a multidimensional variable describing identity, norms and relations between individuals^7^, can influence access to testing, diagnosis and medical care, and significantly impact the availability of social, economical and logistic support^6^. Gender can also influence preventative and risk behavior, possibly impacting the course of the infection.

Several calls urging the inclusion of sex and gender into COVID-19 trials have been published^6,8,9^. Excluding one sex from clinical trials and omitting to disaggregate results by sex can lead to an increased incidence of unwanted side effects in the untested population^10^ due to overmedication and other factors^11^. Not addressing the gender dimension hampers the opportunity to reduce inequality in healthcare, promote preventative action and modulate the course of the infection^12^. Given these potential risks for the health of a large fraction of the infected population, we investigated the consideration of sex and/or gender as an analytical variable in currently registered trials for SARS-CoV-2/COVID-19.

## Results

We identified 2,484 registered SARS-CoV-2/COVID-19 trials. 1,081 trials are observational (924, and a further 157 patient registry trials) and 1,381 interventional (for further specifications see Data and Methods). 70% of the observational trials and 55% of the interventional trials are single-facility studies. 514 of the trials have at least one location in the United States, 367 in France, 134 in Italy, 125 in China and 121 in Spain (Figure 1).

Of the registered trials, 416 (16.7%) mention sex/gender as recruitment criterion/recorded variable, while 103 (4.1%) allude to sex/gender in the description of the analysis phase. 72 studies (2.9%) focus solely on one sex (Table 1); 61 recruiting only female participants and 11 only male ones. Female-only studies mostly focus on the relation between COVID-19 and pregnancy outcomes. One trial explicitly addresses the impact of COVID-19 on the transgender population. The remainder of the trials (76.2%) do not address sex/gender in the study protocol registration on http://ClinicalTrials.gov, beyond the mandated selection from a list of the eligible groups for the study (‘Male’, ‘Female’, or ‘All’). Of the 2,484 trials, 191 (7.7%) trials mention gender in the trial documentation. Attention to sex/gender at the trial design and registration phase is generally low. The trial protocols that consider sex/gender in any form upon recruitment are mostly interventional (70.4%) and the trials that consider sex/gender as a variable upon analysis are mostly observational (70.9%). Only 20 of the 1381 interventional trials plan to consider sex as a variable upon analysis.

**Table 1.**
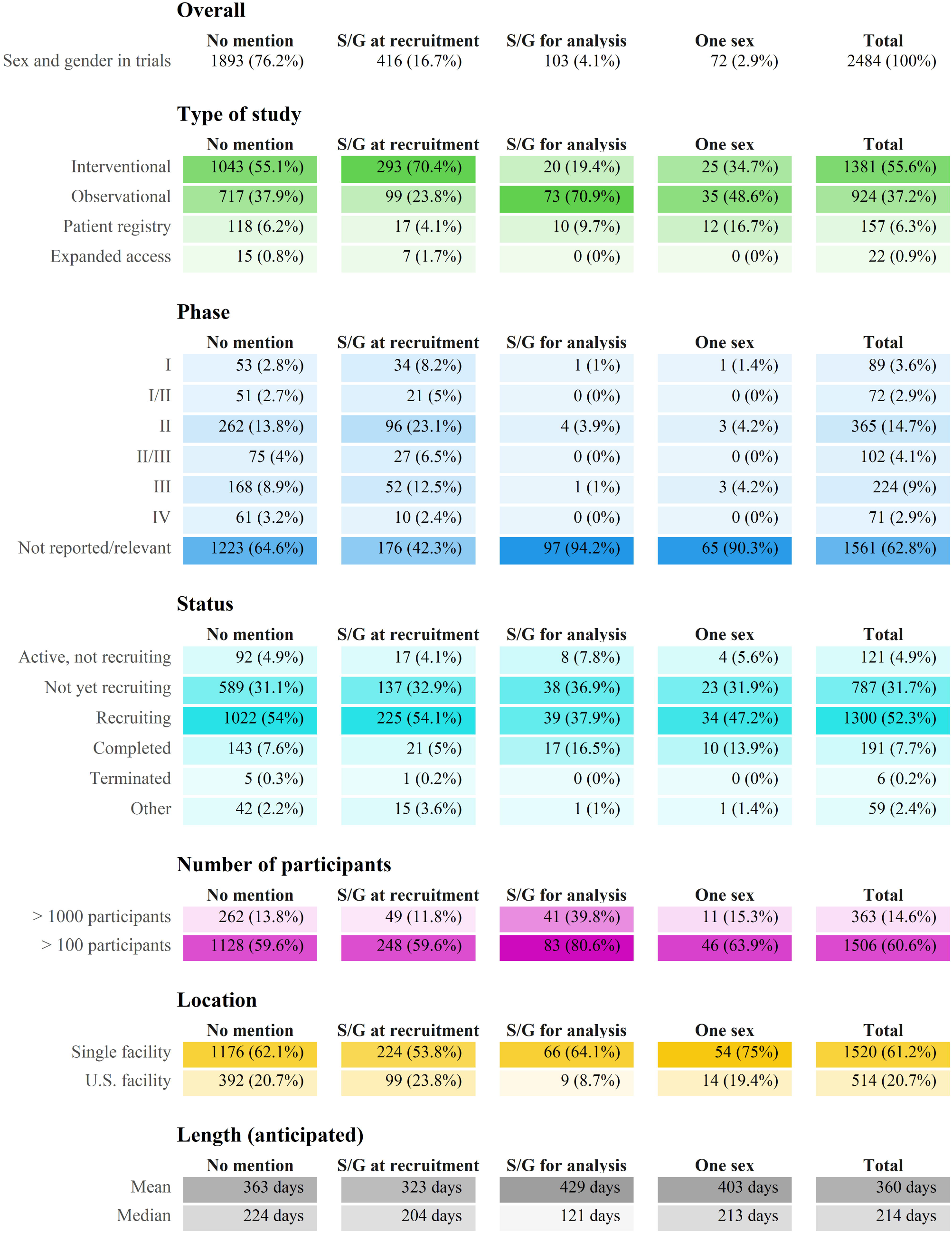
Characteristics of the included trials

|  | No mention | S/G at recruitment | S/G for analysis | One sex | Total |
| --- | --- | --- | --- | --- | --- |
| Sex and gender in trials | 1893 (76.2%) | 416 (16.7%) | 103 (4.1%) | 72 (2.9%) | 2484 (100%) |

Type of study
|  | No mention | S/G at recruitment | S/G for analysis | One sex | Total |
| --- | --- | --- | --- | --- | --- |
| Interventional | 1043 (55.1%) | 293 (70.4%) | 20 (19.4%) | 25 (34.7%) | 1381 (55.6%) |
| Observational | 717 (37.9%) | 99 (23.8%) | 73 (70.9%) | 35 (48.6%) | 924 (37.2%) |
| Patient registry | 118 (6.2%) | 17 (4.1%) | 10 (9.7%) | 12 (16.7%) | 157 (6.3%) |
| Expanded access | 15 (0.8%) | 7 (1.7%) | 0 (0%) | 0 (0%) | 22 (0.9%) |

Phase
|  | No mention | S/G at recruitment | S/G for analysis | One sex | Total |
| --- | --- | --- | --- | --- | --- |
| I | 53 (2.8%) | 34 (8.2%) | 1 (1%) | 1 (1.4%) | 89 (3.6%) |
| I/II | 51 (2.7%) | 21 (5%) | 0 (0%) | 0 (0%) | 72 (2.9%) |
| II | 262 (13.8%) | 96 (23.1%) | 4 (3.9%) | 3 (4.2%) | 365 (14.7%) |
| II/III | 75 (4%) | 27 (6.5%) | 0 (0%) | 0 (0%) | 102 (4.1%) |
| III | 168 (8.9%) | 52 (12.5%) | 1 (1%) | 3 (4.2%) | 224 (9%) |
| IV | 61 (3.2%) | 10 (2.4%) | 0 (0%) | 0 (0%) | 71 (2.9%) |
| Not reported/relevant | 1223 (64.6%) | 176 (42.3%) | 97 (94.2%) | 65 (90.3%) | 1561 (62.8%) |

Status
|  | No mention | S/G at recruitment | S/G for analysis | One sex | Total |
| --- | --- | --- | --- | --- | --- |
| Active, not recruiting | 92 (4.9%) | 17 (4.1%) | 8 (7.8%) | 4 (5.6%) | 121 (4.9%) |
| Not yet recruiting | 589 (31.1%) | 137 (32.9%) | 38 (36.9%) | 23 (31.9%) | 787 (31.7%) |
| Recruiting | 1022 (54%) | 225 (54.1%) | 39 (37.9%) | 34 (47.2%) | 1300 (52.3%) |
| Completed | 143 (7.6%) | 21 (5%) | 17 (16.5%) | 10 (13.9%) | 191 (7.7%) |
| Terminated | 5 (0.3%) | 1 (0.2%) | 0 (0%) | 0 (0%) | 6 (0.2%) |
| Other | 42 (2.2%) | 15 (3.6%) | 1 (1%) | 1 (1.4%) | 59 (2.4%) |

Number of participants
|  | No mention | S/G at recruitment | S/G for analysis | One sex | Total |
| --- | --- | --- | --- | --- | --- |
| > 1000 participants | 262 (13.8%) | 49 (11.8%) | 41 (39.8%) | 11 (15.3%) | 363 (14.6%) |
| > 100 participants | 1128 (59.6%) | 248 (59.6%) | 83 (80.6%) | 46 (63.9%) | 1506 (60.6%) |

Location
|  | No mention | S/G at recruitment | S/G for analysis | One sex | Total |
| --- | --- | --- | --- | --- | --- |
| Single facility | 1176 (62.1%) | 224 (53.8%) | 66 (64.1%) | 54 (75%) | 1520 (61.2%) |
| U.S. facility | 392 (20.7%) | 99 (23.8%) | 9 (8.7%) | 14 (19.4%) | 514 (20.7%) |

Length (anticipated)
|  | No mention | S/G at recruitment | S/G for analysis | One sex | Total |
| --- | --- | --- | --- | --- | --- |
| Mean | 363 days | 323 days | 429 days | 403 days | 360 days |
| Median | 224 days | 204 days | 121 days | 213 days | 214 days |

Footer: Number of participants and length are mostly anticipated values. Only 83% of the registered trials record facility location. Abbreviation: S/G – sex/gender

The PubMed search retrieved 11 original reports of randomized controlled trials. All of these reported the number of female and male participants (44.6% of subjects were female), while none reported explicit details about the inclusion of sex upon analysis, nor disaggregated results by sex.

## Discussion

Although sex appears to be an important determinant of mortality risk and immunologic responses to COVID-19^5^, currently registered clinical trials mostly omit sex as a specific recruitment or analytical criterion. None of the identified publications of randomized-controlled trials report sex-specific analyses nor do they disaggregate results by sex. This suggests that a lack of consideration of sex and/or gender upon trial registration will not be revised during trial execution, leading to a significant information gap.

Gender is given even less consideration. Very few studies address gender as a recruitment or analytical variable. This applies equally to observational and interventional trials. Gender is, however, a relevant risk factor. For example, recent reports highlight that infected healthcare workers worldwide are more frequently women^13^ and that women might be more affected by persistent symptoms after an initial COVID-19 infection^14^.

Some aspects need to be taken into consideration when looking at our findings. First, the size of the trial might impact the inclusion of sex as an analytical variable. Interventional trials in the sample tended to be smaller, which might limit the ability of experimenters to disaggregate analyses while maintaining statistical power. Second, disciplinary culture might affect the decision to perform sex and gender-sensitive analysis. For example, researchers conducting large observational trials might be trained to consider a vast array of social determinants, which biomedical researchers might not prioritize in interventional trials. Third, national requirements for trial performance might play a role in the decision to include sex and/or gender as analytical variables. We employed the US-based database “http://ClinicalTrials.gov”; this will favor the inclusion of US-based trials, as seen in our results. The NIH policies and recommendations^15^ might prompt a higher degree of awareness for sex-sensitivity in the trials registered in the US. Similar effects might be expected in other countries with comparable policies.

In light of the current results, we urge researchers working in the field of SARS-CoV2 and COVID-19 to systematically apply a sex-specific methodology. This entails: (a) The recording of sex of all participants; (b) The inclusion of sex as an independent variable into multivariate analysis; (c) The performance of sex-disaggregated analyses; (d) Results should be reported in a sex-disaggregated manner to allow for unambiguous identification of differences in effectiveness, side effects and mortality. Availability of disaggregated data will also facilitate sex-specific meta-analyses in the future; (e) Interaction with other factors, such as age, co-morbidity, exposure, susceptibility and others should be considered, if applicable.

While gender-sensitive research is still defining its methodological gold standards for the (bio)medical field^16^, resources for sex-specific analysis are available in multiple formats, ranging from scientific publications^17^, to websites and videos^18-20^. Sex-specific analysis is not an exceedingly complex process; the lack of its systematic performance might be due to lack of awareness more than being a deliberate decision. The presented data should be considered an alarming signal, which can still be addressed. Sex and gender should be systematically considered in COVID-19 trial design to guarantee safe and efficacious therapeutic options for all patients.

## Methods

We queried the AACT “Aggregate Analysis of http://ClincalTrials.gov” (https://www.ctticlinicaltrials.org/aact-database) relational database, which contains all publicly available http://ClinicalTrials.gov data, on June 6, 2020 and on July 7, 2020.

The official trial title, brief summary, detailed description and conditions (being studied) fields were searched for variations of: *Coronavirus, SARS-CoV-2, Covid-19* and *2019 nCoV*. We included all trials submitted to http://ClinicalTrials.gov in 2020. Trials without a search term match in the title or conditions were briefly inspected and excluded if SARS-CoV-2/COVID-19 was not the focus of the trial (e.g. just mentioned as reason for interruption of recruitment). Almost 98% of our final sample has one of the search terms in the official trial title or in the conditions.

We identified attention to sex/gender in the trial design phase by searching the study protocol registration fields – title, brief summary, detailed description, eligibility criteria descriptions and (primary, secondary, other) outcome measures – for the following terms (and their plurals): *sex, gender, woman, female, man, male, girl, boy, pregnan*, HRT, hormone replacement therapy, estrogen, progesterone, testosterone and transg\**. Two independent coders (EB and SOP) analyzed all identified trials according to three categories: a) sex/gender as an analysis criterion, b) sex/gender only mentioned in the context of a recruitment criterion or sex/gender of participants is recorded, and c) spurious match/no relevant sex/gender mention.

For inclusion into category a) (analysis criterion) we looked for evidence giving a reasonable expectation of an intention to include sex/gender as an analytical variable. Statements such as that treatment groups would be sex-matched to controls, that results would be stratified by sex etc. were considered permissible and no further statistical details were expected. If sex/gender representation and differences were addressed in an introduction or literature summary, but no mention was made in the outline of the data analysis protocol, we did not consider that sufficient. Similarly if ‘demographic variables’ were identified as part of the analytical variables set, but sex/gender was not explicitly mentioned in that particular discussion, we did not include that trial.

Trials in category b) (recruitment criterion) paid attention to sex i.e in the form of stated intent to at least record/report the sex of participants, or an explicit recruitment statement covering both sexes/genders. Here, the sole mentioning of “all” in the predefined “sexes eligible for study” section was not considered sufficient, as this registration step does not represent a marker of specific focus on the topic. Likewise, a focus on the sex of donors or parents, but not the recipients or children, who were the focus of the study, was not considered sufficient for inclusion. Phrasing such as ‘either/any sex/gender’ or that recruiting was ‘irrespective of sex/gender’ was not considered a strong enough indicator of a focus on recruiting a gender diverse sample.

Trials included in category c) (no match) either had no match to one of our sex-search terms (the vast majority), or were spurious sex matches. The latter set included trials where, for example, the only mention to males/females was in the context of contraception requirements, pregnancy tests etc.

We searched the PubMed database on June 2, 2020 for publications on SARS-CoV-2/COVID-19 trials using the following search terms: “covid-19”[tw] AND trial[ti]. We only analyzed original publications of randomized controlled trials. Two independent coders (MWN and SOP) evaluated the degree of sex/gender-specific analysis in the selected publications. Sex/gender-specific analysis could range from a) sole mention of participating numbers of women/men in the trial, b) explicit incorporation of sex/gender as analytical variable, c) reporting of sex/gender-disaggregated analyses.

## Data Availability

Data is available upon reasonable request from the authors and will be made openly available upon publication.

## Author contributions

SOP, MWN and JPA developed the idea, EB, MWN and SOP conducted the analyses, EB and SOP wrote the paper, all authors reviewed the paper for important intellectual content.

All authors have seen and approved this final version of the manuscript.

